# Age-dependent impairment in antibody responses elicited by a homologous CoronaVac booster dose

**DOI:** 10.1101/2022.10.04.22280704

**Authors:** Bruno Andraus Filardi, Valter Silva Monteiro, Pedro Vellosa Schwartzmann, Vivian do Prado Martins, Luis Eduardo Rosa Zucca, Gabriela Crispim Baiocchi, Anne M. Hahn, Nicholas F. G. Chen, Kien Pham, Eddy Pérez-Then, Marija Miric, Vivian Brache, Leila Cochon, Rafael A. Larocca, Yale SARS-CoV-2 Genomic Surveillance Initiative, Roberto Della Rosa Mendez, Douglas Bardini Silveira, Aguinaldo Roberto Pinto, Julio Croda, Inci Yildirim, Saad B. Omer, Albert I. Ko, Sten H. Vermund, Nathan D. Grubaugh, Akiko Iwasaki, Carolina Lucas

## Abstract

The emergence of the SARS-CoV-2 Omicron sublineages resulted in drastically increased transmission rates and reduced protection from vaccine-induced immunity. To counteract these effects, multiple booster strategies were used in different countries, although data comparing their efficiency in improving protective immunity remains sparse, especially among vulnerable populations, including older adults. The inactivated CoronaVac vaccine was among the most widely distributed worldwide, particularly in China, and South America. However, whether homologous *versus* heterologous booster doses in those fully vaccinated with CoronaVac induce distinct humoral responses and whether these responses vary across age groups remain unknown. We analyzed plasma antibody responses from CoronaVac-vaccinated younger or older individuals in central and south America that received a homologous CoronaVac or heterologous BNT162b2 or ChAdOx1 booster vaccines. We found that both IgG levels against SARS-CoV-2 spike or RBD, as well as neutralization titers against Omicron sublineages, were substantially reduced in participants that received homologous CoronaVac when compared to heterologous BNT162b2 or ChAdOx1 booster. This effect was specifically prominent in recipients older than 50 years of age. In this group, CoronaVac booster induced low virus-specific IgG levels and failed to elevate their neutralization titers against any omicron sublineage. Our results point to significant inefficiency in mounting protective anti-viral humoral immunity in those who were primed with CoronaVac followed by CoronaVac booster, particularly among older adults, urging a heterologous regimen in high-risk populations fully vaccinated with CoronaVac.

**One Sentence Summary:** Homologous CoronaVac boosters do not improve neutralization responses against current VOCs in older adults in contrast to heterologous regimens.

## INTRODUCTION

The emergence of the SARS-CoV-2 Omicron variant was a turning point in the COVID-19 pandemic. Within 2 months of its first report, in November 2021, the BA.1 sublineage became the dominant variant worldwide, followed by the appearance of several additional sublineages. Omicron BA.1 was first displaced by the BA.2 sublineage, subsequentially followed by its descendants BA.2.12.1, BA.4, and BA.5 (*1-3*). While specific amino acid changes are shared between all Omicron sublineages, a substantial level of mutations between them was reported in the spike receptor binding domain region (RBD). As a result, these antigenic differences of Omicron sublineages drastically reduced their susceptibility to vaccine-induced neutralizing antibodies, increasing the need for additional vaccination boosters, especially among vulnerable groups (*2-6*). Current COVID-19 vaccines, including Pfizer-BioNTech, ChAdOx1 nCoV-19, and CoronaVac are highly effective against hospitalization and death caused by SARS-Cov-2, despite their different formulation (*7-9*). Whereas the Pfizer/BioNTech BNT162b1 vaccine is based on mRNA encoding the complete stabilized S protein encapsulated within lipid nanoparticles (LNP) (*10*), the ChAdOx1 nCoV-19 vaccine consists of a replication-deficient chimpanzee adenoviral vector ChAdOx1 encoding the SARS-CoV-2 spike protein (ChAdOx1-S)(*11*); and the CoronaVac is a whole-virus β-propiolactone-inactivated vaccine with aluminum hydroxide-adjuvant, authorized for use in 48 countries (*12*).

Brazil is the second country in the world in terms of the absolute number of COVID-19-related deaths, with almost 700,000 fatalities to date. Like many other countries, Brazil widely administered CoronaVac, which helped control the number of SARS-CoV-2-related hospitalizations and deaths, especially at the beginning of the pandemic (*13, 14*). However, as SARS-CoV-2 Omicron sublineages emerged with increased transmissibility and extensive escape from previously established immunity, several concerns were raised regarding reduced vaccine effectiveness, particularly for vulnerable populations with suboptimal immunity. Thus, all three vaccines, Pfizer-BioNTech, ChAdOx1 nCoV-19, and CoronaVac, have been administered as booster doses to address both waning immunity and reduced effectiveness against SARS-CoV-2 variants in Brazil and elsewhere (*11, 14-16*). However, despite its potential significance, given that CoronaVac remains among the top distributed vaccines, studies comparing humoral immune responses from patients primed with CoronaVac followed by boosters with distinct vaccine formulations are largely missing. In addition, studies comparing homologous and heterologous booster against omicron sublineages in high-risk, elderly populations are limited (*17, 18*).

## RESULTS

### Study Cohort

To evaluate virus-specific immune responses following different booster vaccines across age groups and to assess the potential risk of vaccine immune evasion by Omicron infection, we assembled a cohort of CoronaVac-vaccinated individuals that received a homologous CoronaVac or heterologous (BNT162b2 or ChAdOx1) booster vaccine. We investigated antibody titers and vaccine-induced neutralizing responses against the ancestral strain, USA-WA1/2020, B.1.617.2 (Delta variant), BA.1 (Omicron variant) as well as the omicron sublineages, BA.2.12.1, XAF (BA.1/BA.2 circulating recombinant) and BA.5 and compared across age groups. We studied 293 non-hospitalized adult participants who received two doses of CoronaVac vaccine between November 27, 2021 to February 3, 2022, before and after the BNT162b2, ChAdOx1 or CoronaVac booster dose. Plasma samples were collected at baseline (prior to booster) and 28 days after booster (third dose) administration (Fig. 1A) and immunogenicity endpoints included enzyme-linked immunosorbent assays (ELISA) and neutralization assays using the authentic virus. Blood samples were collected at the Serviço de Tratamento ao Câncer de Ribeirão Preto, São Paulo, Brazil. Data from a previously analyzed cohort, composed of participants from the Dominican Republic that received two doses of CoronaVac followed by BNT162b2 booster, was used as a reference (*16*). Study groups were stratified by age (younger adults, <50 years and older adults ≥ 50 years), biological sex, and booster vaccine type. The mean age of the participants was 39.3 ± 15.9, the majority of them females (∼61%). Participants did not significantly differ with respect to their previous SARS-CoV-2 infection status (i.e., previously infected vs. previously uninfected). Basic demographics information, vaccination, and previous infection status are summarized in the Table S1.

**Figure 1.**
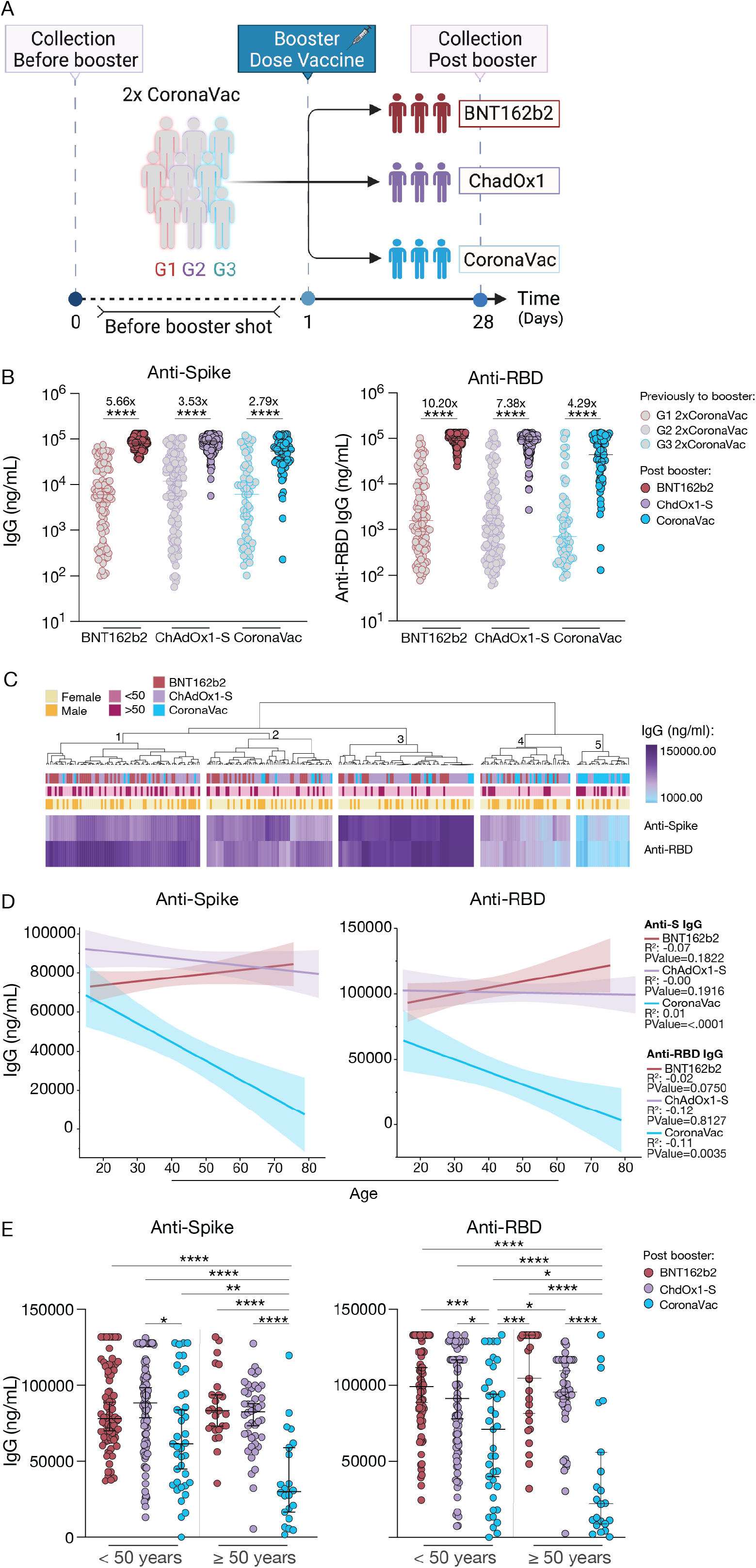
Sars-CoV-2 specific antibodies responses by age post heterologous or homologous booster in individuals fully vaccinated with CoronaVac. **a**, Cohort timeline overview indicated by days after SARS-CoV-2 vaccination booster in CoronaVac previously vaccinated cohort. Participants received boosters of BNT162b2, ChAdOx1-S, or CoronaVac vaccine and plasma samples were collected as indicated. Baseline time point 0, before vaccination booster; and 28 days after the booster dose; participants were stratified by booster dose received in three groups (G1, BNT162b2; G2, ChAdOx1-S; G3, CoronaVac). Created with BioRender. **b**, Plasma reactivity IgG to S protein and RBD measured before and post booster vaccine. S, spike. RBD, receptor binding domain at baseline and after booster dose. The median fold change values in antibody titer post booster vaccination are indicated. **c**, Heat map comparison of plasma Anti-S and Anti-RBD levels within participants CoronaVac primed followed by BNT162b2, ChAdOx1-S, or CoronaVac. Subjects are arranged across rows, and color intensity indicates anti-virus IgG concentration. Analyses were performed 28 days post booster dose. **d**, Correlation and linear regression comparisons of virus-specific Anti-S and Anti-RBD IgG levels by age post vaccination booster. Regression lines are shown as red (BNT162b2 booster), purple (ChAdOx1-S booster) or blue (CoronaVac booster) Pearson’s correlation coefficients and linear regression significance are colored accordingly; shading represents 95% confidence interval. **e**, Plasma reactivity to S protein and RBD measured by ELISA at 28 days post booster by age groups. < 50 years, vaccinated participants aged 49 or younger; ≥ 50 years, vaccinated participants aged 50 years or older. Significance was assessed by One-way ANOVA corrected for multiple comparisons using Tukey’s method. Previously to booster dose: G1 (n = 101), G2 (n = 131), and G3 (n = 61). Post booster dose: BNT162b2 (n = 101), ChAdOx1-S (n = 131), and CoronaVac (n = 61). Each dot represents a single individual. Horizontal bars represent average ± s.d. ****p < .0001 ***p < .001 **p < .01 *p < .05.

### Defective age-associated SARS-CoV-2 antibody response post CoronaVac homologous, but not heterologous vaccination-booster regimen

Plasma antibody reactivity to Spike protein (S) and receptor binding domain (RBD) of SARS-CoV-2 were measured at baseline, and 28 days post COVID vaccines booster (Fig. 1A). Levels of IgG RBD-specific binding antibodies were similar at the baseline for the Brazilian and Dominican Republic cohorts, although a slight, but significant, reduction in virus-specific IgG levels against spike protein was observed for the Dominican Republic participants (Fig. S1A). As expected, at 28 days post booster dose we detected a significant increase in the virus-specific IgG titers for all groups (Fig.1B). However, individuals that received the homologous regimen, CoronaVac prime followed by CoronaVac booster, mounted lower anti-S and anti-RBD titers compared to individuals that received a heterologous regimen (Fig.1B, Fig. S1B). No difference was observed among participants that received CoronaVac prime followed by BNT162b2 or ChAdOx1-S booster regimen (Fig. S1B). Consistently, an unsupervised heatmap assembled using virus-specific antibody titers and the main cohort demographics revealed marked changes in CoronaVac/CoronaVac recipients, especially among older participants compared to young adults (Fig. 1C). Five main clusters of vaccinated participants emerged, and the distribution of participants matched the vaccination booster received and correlated with age groups. Cluster 3 primarily comprised younger participants that received CoronaVac/ChAdOx1 regimen with the highest virus-specific IgG titers. Clusters 1 and 2 contained mainly both younger and older adults that received CoronaVac/ChAdOx1 or CoronaVac/BNT162b2 regimen with moderate levels of anti-S and anti-RBD post booster shot. Cluster 4 and 5 comprised participants with the lowest levels of virus-specific IgG antibodies. The majority of CoronaVac/CoronaVac recipients fell into these clusters, including 77% of the participants aged 50 or more from this group (Fig. 1C). Levels of anti-spike and -RBD IgG declined with age for participants that received the homologous regimen in contrast to participants that received a heterologous booster regimen; in fact, the latter did not present an inverse correlation between age and antibody levels post booster (Fig. 1D). No differences were observed in antibody levels between vaccinated participants of different sexes after stratification by age (Fig. S1C).

Further analysis across age groups revealed that the levels of anti-spike and anti-RBD IgGs were reduced in younger adults that received CoronaVac compared to younger groups that received BNT162b2 or ChAdOx1-S booster. Moreover, IgG levels were substantially lower in participants aged 50 or older receiving homologous boosters than in the younger adult groups or older adult participants with a heterologous regimen (Fig. 1E). No differences were observed in antibody levels between vaccinated participants that received BNT162b2 or ChAdOx1-S booster after stratification by age (Fig. 1E). These observations point to significant inefficiency in mounting virus-specific antibodies in participants primed with CoronaVac followed by CoronaVac booster, particularly among participants older than 50 years old. Indeed, BNT162b2 and ChAdOx1-S booster were associated with higher vaccine antibody-induction as compared to CoronaVac in both younger or over-50 group, resulting in a 10.3/5.8- fold increase (<50 years) and 9.9/14.6-fold increase (>50 years) in anti-RBD IgG titers, compared to 4.2-fold increase (<50 years) and 7.0-fold increase (>50 years) for CoronaVac booster recipients (Fig. S1D). Together, these data indicate that participants that received a homologous regimen, CoronaVac prime followed by CoronaVac booster, develop lower virus-specific antibody responses with a remarkable impact in older participants. Of note, these differences appear to be vaccine-specific, rather than resulting from an aging-associated impaired immune response.

### Homologous CoronaVac boosters do not improve neutralization responses against Omicron sub-lineages in older adults

A central premise for current COVID-19 vaccine programs is that neutralizing antibodies correlate with protection against SARS-CoV-2 infection (*19, 20*). Given our observation that CoronaVac primed/booster participants aged 50 years or more had lower antibodies compared to other analyzed groups, we hypothesized that this could lead to sub-protective neutralizing responses. We next assessed whether different booster strategies impacted also neutralizing responses by measuring plasma neutralization activity longitudinally using half-maximal plaque reduction neutralizing assays (PRNT50) against circulating SARS-CoV2-authentic isolates, including lineage A (ancestral strain, USA-WA1/2020), B.1.617.2 (Delta variant), BA.1 (Omicron variant) as well as the omicron sublineages, BA.2.12.1, XAF (BA.1/BA.2 circulating recombinant) and BA.5. Individuals who were fully vaccinated with CoronaVac and received BNT162b2 booster displayed the highest increase in neutralization activity (10.8-, 7.0-, 3.5-, 5.4-, 3.8- and 3.6-fold increase) against ancestral, delta, BA.1, BA.2.12.1, XAF and BA.5 respectively, 28 days after the booster shot (Fig. 2A and Fig. S2A). The BNT162b2 group was followed by the participants that were fully vaccinated with CoronaVac and received ChAdOx1 booster, which showed an increase in neutralization activity (3.4-, 6.5-, 2.2-, 2.4-, 2.7- and 2.1-fold) against ancestral, delta, BA.1, BA.2.12.1, XAF and BA.5, respectively. After booster, CoronaVac/CoronaVac recipients developed neutralizing antibody titers against ancestral strain, Delta, BA.2.12.1 and XAF variants (2.6-, 3.4-,1.2- and 1.9 fold increase, respectively). Despite this increase, PRNT50 values were significantly reduced compared to levels induced by the participants that received a heterologous booster. Additionally, no statistical differences were observed after booster for CoronaVac/CoronaVac recipients upon assessing neutralizing antibodies titers against BA.1 and BA.5 (Fig. 2A and Fig. S2A).

**Figure 2.**
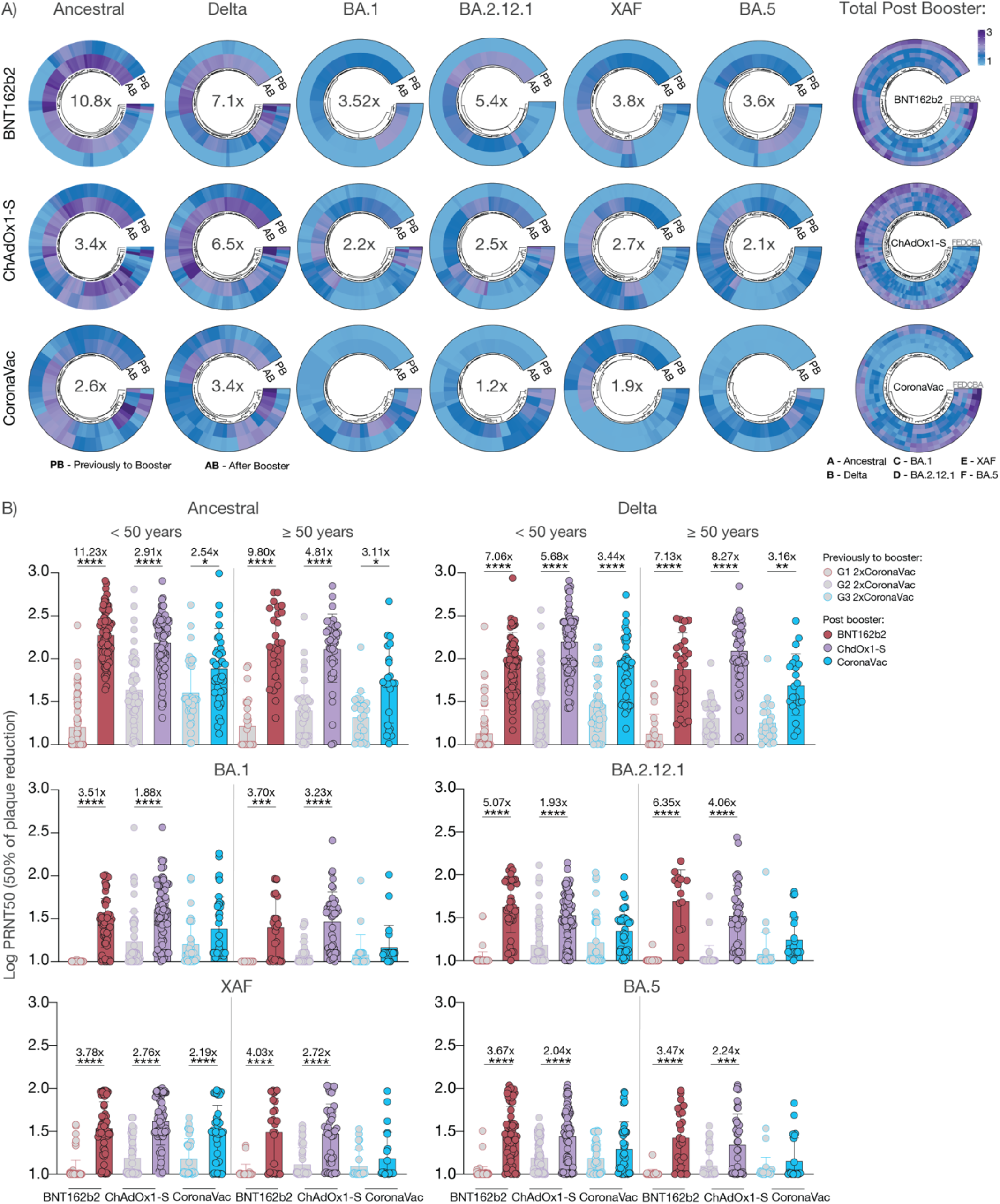
Neutralization responses following heterologous or homologous booster in individuals fully vaccinated with CoronaVac. **a-b**, Plasma neutralization capacity against the ancestral strain (WA1, USA), Delta, BA.1, BA.2.12.1, XAF, and BA.5 before and after booster dose. Participants received boosters doses of BNT162b2, ChAdOx1-S, or CoronaVac. **a**, Circular heat map comparisons of neutralization titers from fully vaccinated CoronaVac participants at baseline and 28 days post booster dose. PB, previously to booster dose. AB, after booster dose. Color intensity indicates log PRNT50 for each specific SARS-CoV-2 variant tested. PRNT50, 50% f plaque reduction. The median fold change values in neutralization titers post booster vaccination are indicated inside the circles. Total neutralization titers against: ancestral strain, Delta and BA.1 (BNT162b2, n = 101; ChAdOx1-S, n = 131; CoronaVac, n = 61); BA.2.12.1(BNT162b2, n = 49; ChAdOx1-S, n = 131; CoronaVac, n = 61); XAF, and BA.5 (BNT162b2, n = 86; ChAdOx1-S, n = 131; CoronaVac, n = 61).**b**, Plasma neutralization capacity comparisons by age groups. < 50 years, vaccinated participants aged 49 or younger; ≥ 50 years, vaccinated participants aged 50 years or older. The median fold change values in neutralization titers post booster vaccination are indicated. Neutralization titers for < 50 years participants against: ancestral strain, Delta and BA.1 (BNT162b2, n = 76; ChAdOx1-S, n = 87; CoronaVac, n = 39); BA.2.12.1(BNT162b2, n = 35; ChAdOx1-S, n =87; CoronaVac, n = 39); XAF, and BA.5 (BNT162b2, n = 35; ChAdOx1-S, n = 87; CoronaVac, n = 39). Neutralization titers for ≥ 50 years participants against: ancestral strain, Delta and BA.1 (BNT162b2, n = 25; ChAdOx1-S, n = 44; CoronaVac, n = 22); BA.2.12.1(BNT162b2, n = 14; ChAdOx1-S, n =44; CoronaVac, n = 22); XAF, and BA.5 (BNT162b2, n = 25; ChAdOx1-S, n = 87; CoronaVac, n = 39). Significance was assessed by One-way ANOVA corrected for multiple comparisons using Tukey’s method. Each dot represents a single individual. Horizontal bars represent average ± s.d.****p < .0001 ***p < .001 **p < .01 *p < .05.

Importantly, no statistical differences were observed in neutralization titers prior to the booster shot for the Brazilian cohort (Fig. S2B). Stratification by age revealed that similar to antibody levels, neutralization titers were significantly lower in CoronaVac/CoronaVac recipients of different ages, particularly age 50 or older when compared to participants that received heterologous regimen (Fig. S2C and D). Increased PRNT50 values were observed for all participants, independent of age group, against all variants and sub-lineages tested, that received CoronaVac prime followed by BNT162b2 or ChAdOx1 booster dose (Fig. 2B). Despite an increased in neutralization levels, post booster, against ancestral and delta variant, remarkably, older participants that received the homologous regimen did not show increased neutralization titers against BA.1 or additional omicron sub-lineages post booster dose (Fig. 2B). With exception of neutralizing antibodies against XAF sub-lineage, younger participants that received the homologous CoronaVac regimen also did not show improved neutralization titers against Omicron sub-lineages post booster shot (Fig. 2B). Of note, although previous studies showed that neutralization titers against VOCs were higher for previously infected individuals when compared to non-previously infected individuals, we did not find such correlation in titers against Omicron sub-lineages (Fig. S3A, B, and C). Altogether, these analyses suggest that administration of heterologous vaccine boosters to people fully vaccinated with CoronaVac significantly improves their humoral responses, including neutralization titers against currently circulating omicron VOCs. Additionally, these data suggest that homologous CoronaVac boosters do not significantly improve antibody responses against Omicron sub-lineages, particularly in the older population.

## DISCUSSION

Safe and effective vaccines against SARS-CoV-2 are essential resources to manage the ongoing pandemic. In Brazil, where more than 680,000 COVID-related deaths place the country as number 2 in the world, reduced speed in vaccine distribution rather than hesitancy, has been associated with the increased fatality rate in the earlier phases of the pandemic. As in China and several countries in South and Central America, the inactivated COVID vaccine, CoronaVac was widely distributed in the early phases in the pandemic. Therefore, even with the declining fatality rate during the Omicron wave, it is crucial to design booster strategies that continue to induce protective immunity, particularly in the vulnerable population. In this study, we found that a heterologous vaccine regimen, composed of a two-dose CoronaVac prime followed by a single BNT162b2 or ChAdOx1 booster, induced higher levels of neutralizing and virus-specific antibodies against VOCs when compared to a homologous CoronaVac/CoronaVac regimen.

Age-related decreases in vaccine-induced antibodies and cellular responses were reported by several studies, in those primed with mRNA, viral vectors, or inactivated vaccines. In older populations, lower IgG and IgA anti-S and RBD titers were observed as well as neutralization responses, with notable reductions in participants aged 80 or more post initial vaccination doses (*17, 18, 21-23*). In contrast to the mRNA BNT162b2, or the viral vector ChAdOx1vaccine booster, we observed an inverse correlation between age and virus-specific antibody responses in recipients of the CoronaVac booster, suggesting that formulation of vaccine boosters may dictate or reinforce age-specific decline in immunity. Differences in antibody responses upon heterologous versus homologous strategies were more pronounced in neutralizing responses against omicron sub-lineages, especially among the participants over 50 years old. Previous studies have reported increased anti-SARS-CoV-2 spike antibodies, and neutralization titers against ancestral, delta, and BA.1 by heterologous regimen compared to homologous vaccination (*14, 15, 24, 25*). Our findings extend these observations for omicron sub-lineages and caution against the use of CoronaVac as a booster strategy. In addition, our analysis supports real-world vaccine efficacy data reporting reduced effectiveness, increased hospitalization or death rates of CoronaVac homologous booster strategy in elderly when compared to heterologous booster (51 vs 78%, respectively) (*26*). Vaccine- or age-related mechanisms underlying these differences remain to be identified. Our study did not examine cellular immune responses, mucosal immune responses, or the durability of booster-induced immunity. Nevertheless, our findings provide a better understanding of vaccine-induced humoral responses for vulnerable groups during the omicron wave and could potentially guide future immunization strategies and public health policies.

## Data Availability

All the background information of participants and data generated in this study are included in Source Data Figure1. The genome information and aligned consensus genomes for SARS-CoV-2 variants used in this study are available on NCBI (GenBank Accession numbers: ancestral lineage A MZ468053, Delta MZ468047, Omicron (BA.1) OL965559, Omicron (BA.2.12.1) ON411581, Omicron (XAF) OP031604, Omicron (BA.5) OP031606). Additional correspondence and requests for materials should be addressed to the corresponding authors.

## Acknowledgements

We are very grateful to all study participants who kindly donated specimens for this study. We thank M. Linehan for technical and logistical assistance. We also would like to thank D. Mucida who provided insight and expertise that greatly assisted the data analysis and comments that significantly improved this manuscript. Brazilian team is grateful to Marco Krieger (Vice-president of FioCruz), Bárbara Furtado, Victor Bertollo Gomes Porto, Programa Nacional de Imunizaçā Estefania Caires, SECOVID, Multiplan, Instituto do Cancer Brasil de Ensino e Pesquisa, CAPED Ribeirão Shopping, Mariana Kobayashi, Ágora Group and Pec. Team for their contributions to the set-up of the study platform. Gessica Galbiati for the pharmaceutical assistance. Luciana Jarduli, Gabriela Faustino, L. Zucca, Carolina Bonafim, Julia Anelli, Leticia Prado and Otavio Gração for technical assistance. The Dominican Republic team is Julia Anelli, Leticia Prado and Otavio Gração for technical assistance. The Dominican Republic team is grateful to Ms Magaly Caram (PROFAMILIA) and Mark Kelly (Laboratorio de Referencia) for their contributions to the set-up of the study platform. The CoronaVac vaccine were provided by the Brazilian Government and ChAdOx1-S vaccine were provided by FioCruz Brazil.

## Funding

AstraZeneca Externally sponsored scientific research program: Brazilian clinical cohort set-up team. Women’s Health Research at Yale Pilot Project Program: AI.

Fast Grant from Emergent Ventures at the Mercatus Center: AI and NDG.

Mathers Foundation, and the Ludwig Family Foundation, the Department of Internal Medicine at the Yale School of Medicine, Yale School of Public Health and the Beatrice Kleinberg Neuwirth Fund

Howard Hughes Medical Institute: AI.

CAPES-YALE fellowship: VSM.

## Author contributions

Conceived the study: BAF, AI, and CL.

Brazilian cohort set up and sample collection: BAF, PVS, VPM, LERZ, and RAL.

Dominican Republic cohort set up and sample collection: EPT, MM, VC, LC, and SV.

Surveilled, detected and performed virus sequencing: AMH, NFGC, KP, and NDG.

Experiments and data collection: VSM, and CL.

Data analysis: VSM, and CL. Heat map analysis: GCB.

Writing – Original draft: BAF, VSM, AI, and CL.

Writing – Review & editing: RDRM, DBS, ARP, JC, IY, SBO, AIK.

Supervised the project: AI and CL

All authors reviewed and approved the manuscript.

**\* The Yale SARS-CoV-2 Genomic Surveillance Initiative**

Chantal B. F. Vogels^7^, Mallery Breban^7^, Tobias R Koch^7^, Chrispin Chaguza^7^, Irina Tikhonova^7^, Christopher Castaldi^24^, Shrikant Mane^24^, Bony De Kumar^24^, David Ferguson^24^, Nicholas Kerantzas^25^, David Peaper^25^, Marie L Landry^25^, Wade Schulz^26^.

^24^Yale Center for Genome Analysis, Yale University, New Haven, CT, 06510, USA

^25^Department of Laboratory Medicine, Yale New Haven Hospital, CT 06510, USA

^26^Center for Outcomes Research and Evaluation, Yale New Haven Hospital, CT 06510, USA

## Competing interests

A.I. serves as a consultant for RIGImmune, Xanadu and Revelar Biotherapeutics. B.A.F. had lecture fees and sponsored travel by AstraZeneca. L.E.R.Z. served in the advisor board for Zodiac and had lecture fees by AstraZeneca, Bayer, Janssen, Astellas. N.D.G. is a consultant for Tempus Labs to develop infectious disease diagnostic assays. All other authors declare no competing interests.

## METHODS

### Ethics statement

This study was approved by the National Research Bioethics Committee of Brazil (CONEP, CAAE 50457721.9.0000.0175) and National Bioethics Committee of the Dominican Republic (CONABIOS). The participants received two doses of the inactivated whole-virion vaccine CoronaVac followed by a single BNT162b2, ChAdOx-S, or CoronaVac booster dose. The interval between the second dose of CoronaVac and booster shot was at least four weeks. The Brazilian cohort received ChAdOx1-S and CoronaVac boosters, that were administrated between November 27, 2021 and February 03, 2022. The Dominican Republic received two doses of CoronaVac followed by the mRNA vaccine BNT162b2 booster, that was administrated between July 30 and August 27, 2021. Informed consent was obtained from all enrolled vaccinated individuals. None of the participants experienced serious adverse effects after vaccination.

### Study design and Participants

One hundred and ninety-two participants from the Brazil and one hundred and one volunteers from the Dominican Republic were followed serially post-vaccination. Plasma samples were collected at baseline (prior to booster, after two doses of CoronaVac), and 28 days after the booster (third dose) administration. Demographic information was aggregated through a systematic review and was used to construct Supplementary Table 1. The clinical data were collected using REDCap (v5.19.15 @2021 Vanderbilt University) software. Blood acquisition was performed and recorded by a separate team. Vaccinated clinical information and time points of collection information was not available until after processing and analyzing raw dat. ELISA and neutralizations were performed blinded. Documented history of prior SARS-CoV-2 infection was confirmed by absence of SARS-CoV-2-specific antibodies, and information of time window post CoronaVac vaccination are available in the Supplementary Table1.

### Plasma isolation and storage

Whole blood was collected in heparinized CPT blood vacutainers (BD; # BDAM362780 or Greiner; REF 455051BR) and kept on gentle agitation until processing. All blood was processed on the day of collection in a single step standardized method. Plasma samples were collected after centrifugation of whole blood at 600 g for 20 min at room temperature (RT) without brake. The undiluted plasma was transferred to 15-ml polypropylene conical tubes, and aliquoted and stored at −80 °C for subsequent shipping and analysis. Plasma samples were sourced from Dominican Republican and Brazilians participants and were shipped to Yale University. The plasma was aliquoted and heat-inactivated at 56°C for 30 min to inactivate complement prior to micro-neutralization.

### SARS-CoV-2 culture

TMPRSS2-VeroE6 kidney epithelial cells were cultured in Dulbecco’s Modified Eagle Medium (DMEM) supplemented with 1% sodium pyruvate (NEAA) and 10% fetal bovine serum (FBS) at 37°C and 5% CO2. The cell line has been tested negative for contamination with mycoplasma. SARS-CoV-2 lineage A (USA-WA1/2020), was obtained from BEI Resources (#NR-52281). Delta and Omicron subvariants, BA.1, BA2.12.1, XAF and BA.5, were sequenced as part of the Yale Genomic Surveillance Initiative’s weekly surveillance program in Connecticut, United States and then isolated from nasopharyngeal specimens, as previously described(*27*). The pelleted virus was then resuspended in PBS and aliquoted for storage at −80°C. Lineage assignments were confirmed using Pangolin (version 3.1.17) and the respective consensus sequences were submitted to NCBI (GenBank accession: ancestral lineage A = MZ468053, Delta = MZ468047, Omicron (BA.1) = OL965559, Omicron (BA.2.12.1) = ON411581, Omicron (XAF, Spike region derived from BA.2) = OP031604, Omicron (BA.5) = OP031606). Viral titers were measured by standard plaque assay using TMPRSS2-VeroE6. Briefly, 300 µl of serial fold virus dilutions were used to infect Vero E6 cells in DMEM supplemented NaHCO3, 4% FBS 0.6% Avicel RC-581. Plaques were resolved at 48h post-infection by fixing in 10% formaldehyde for 1h followed by 0.5% crystal violet in 20% ethanol staining. Plates were rinsed in water to plaques enumeration. All experiments were performed in a biosafety level 3 laboratory with approval from the Yale Environmental Health and Safety office.

### SARS-CoV-2 specific-antibody measurements

ELISAs were performed as previously described (*27*). Briefly, 96-well MaxiSorp plates (Thermo Scientific #442404) were coated with 50 μl/well of recombinant SARS Cov-2 SARS-CoV-2 S protein (ACROBiosystems #SPN-C52H9-100ug), or RBD (ACROBiosystems #SPD-C52H3-100ug) at a concentration of 2 μg/ml in PBS and were incubated overnight at 4°C. The coating buffer was removed, and plates were incubated for 1h at RT with 200 μl of blocking solution (PBS with 0.1% Tween-20, 3% milk powder). Plasma was diluted 1:800 in dilution solution (PBS with 0.1% Tween-20, 1% milk powder) and 100 μl of diluted serum was added for two hours at RT. Human Anti-Spike (SARS-CoV-2 Human Anti-Spike (AM006415) (Active Motif #91351) was serially diluted to generate a standard curve. Plates were washed three times with PBS-T (PBS with 0.1% Tween-20) and 50 μl of HRP anti-Human IgG Antibody (GenScript #A00166, 1:5,000) diluted in dilution solution added to each well for1 h. Plates were developed with 100 μl of TMB Substrate Reagent Set (BD Biosciences #555214) and then read at a wavelength of 450 nm and 570nm.

### Neutralization assay

Sera from vaccinated individuals were heat treated for 30 min at 56°C. Sixfold serially diluted plasma, from 1:10 to 1:2430 were incubated with SARS-CoV-2 variants, for 1 h at 37 °C. The mixture was subsequently incubated with TMPRSS2-VeroE6 in a 12-well plate for 1h, for adsorption. Then, cells were overlayed with MEM supplemented NaHCO3, 4% FBS 0.6% Avicel mixture. Plaques were resolved at 40 h post infection by fixing in 10% formaldehyde for 1 h followed by staining in 0.5% crystal violet. All experiments were performed in parallel with baseline controls sera, in an established viral concentration to generate 60-120 plaques/well.

### Statistical analysis

All analyses were conducted using GraphPad Prism 8.4.3, JMP 15, R and Morpheus web tool. Multiple group comparisons were analyzed by running parametric (ANOVA) statistical tests. Multiple comparisons were corrected using Tukey’s test as indicated in figure legends. Heatmap of the unsupervised hierarchical cluster of anti-S and anti-RBD was created using the open-source software Morpheus (https://software.broadinstitute.org/morpheus/) applying Euclidian distance metric (*28*). Circular heat maps characterizing the unsupervised hierarchical cluster of log PRNT50 by average linkage method were created using circlize and ComplexHeatmap R packages (*29*).

## Data availability

All the background information of participants and data generated in this study are included in Source Data Figure 1. The genome information and aligned consensus genomes for SARS-CoV-2 variants used in this study are available on NCBI (GenBank Accession numbers: ancestral lineage A = MZ468053, Delta = MZ468047, Omicron (BA.1) = OL965559, Omicron (BA.2.12.1) = ON411581, Omicron (XAF) = OP031604, Omicron (BA.5) = OP031606). Additional correspondence and requests for materials should be addressed to the corresponding authors.

## Supplementary Figures Legend

**Figure S1.**
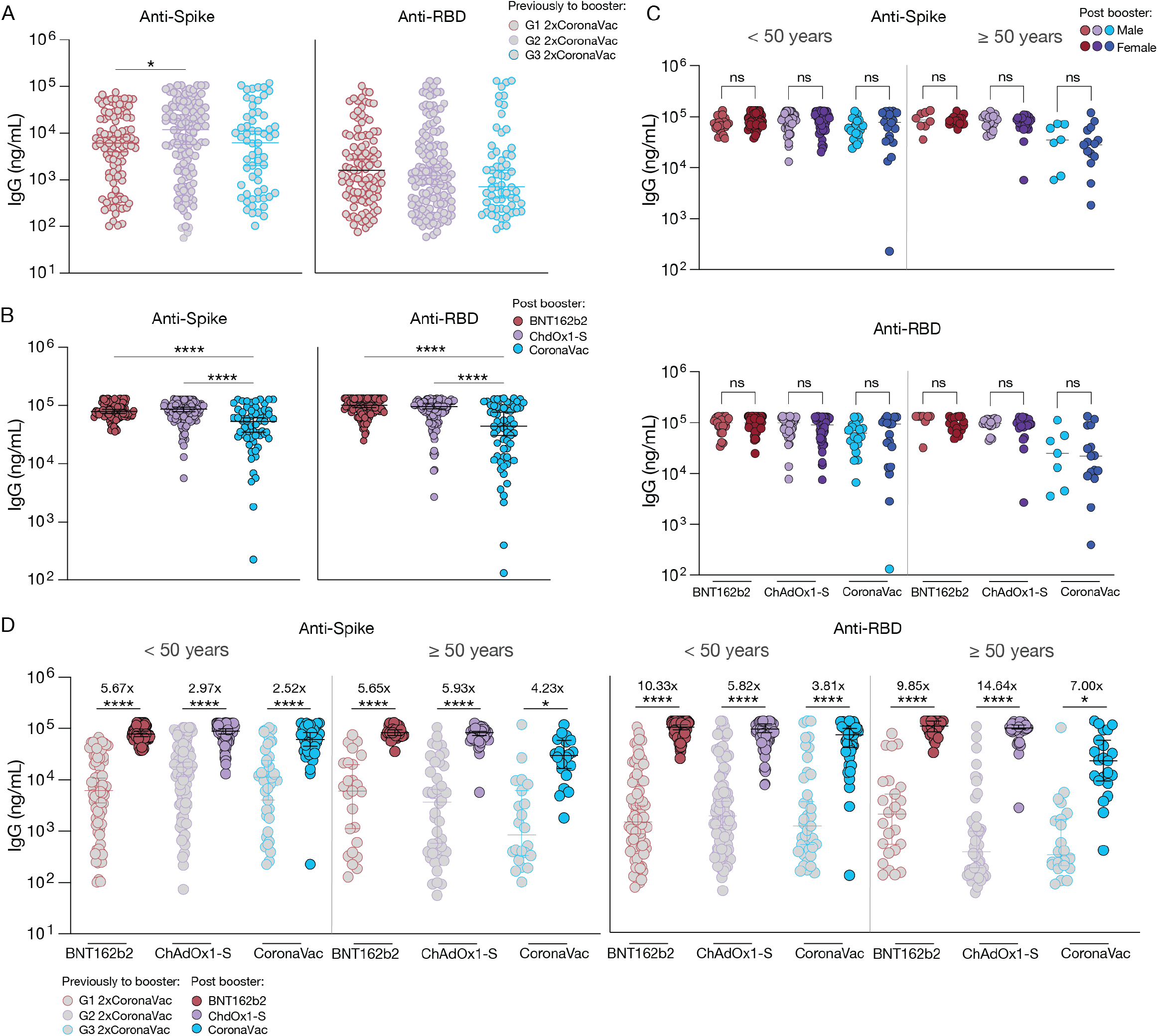
Characterization of Sars-CoV-2 specific antibodies post heterologous versus homologous booster in fully vaccinated CoronaVac cohort. **a, b, c**, and d, Plasma reactivity to S protein and RBD by ELISA. S, spike. RBD, receptor binding domain. Fully vaccinated CoronaVac participants received BNT162b2, ChAdOx1-S, or CoronaVac booster dose. Plasma samples were collected at baseline, prior to vaccination booster and 28 days after the booster dose. Participants were stratified based on the booster dose vaccine into three groups (G1, BNT162b2; G2, ChAdOx1-S; G3, CoronaVac). **a**, IgG virus-specific antibody levels before booster dose. G1 (n = 101), G2 (n = 131), and G3 (n = 61), **b**, IgG virus-specific antibody levels 28 days post booster dose. BNT162b2 (n = 101), ChAdOx1-S (n = 131), and CoronaVac (n = 61). **c**, Anti-S and anti-RBD IgG levels between female and male participants groups at baseline and after booster dose. **d**, Anti-S and anti-RBD IgG levels by age groups at baseline and post booster dose. < 50 years, vaccinated participants aged 49 or younger; ≥ 50 years, vaccinated participants aged 50 years or older. The median fold change values in antibody titer post booster vaccination are indicated. Antibody titers for < 50 years participants: G1, n = 76; G2, n = 55; G3, n = XX; BNT162b2, n = 76; ChAdOx1-S, n = 87; CoronaVac, n = 39). Antibody titers for ≥ 50 years participants: G1, n = XX; G2, n = 38; G3, n = 22; BNT162b2, n = 25; ChAdOx1-S, n = 44; CoronaVac, n = 22. Significance was assessed by One-way ANOVA corrected for multiple comparisons using Tukey’s method. Each dot represents a single individual. Horizontal bars represent average ± s.d. ****p < .0001 ***p < .001 **p < .01 *p < .05.

**Figure S2.**
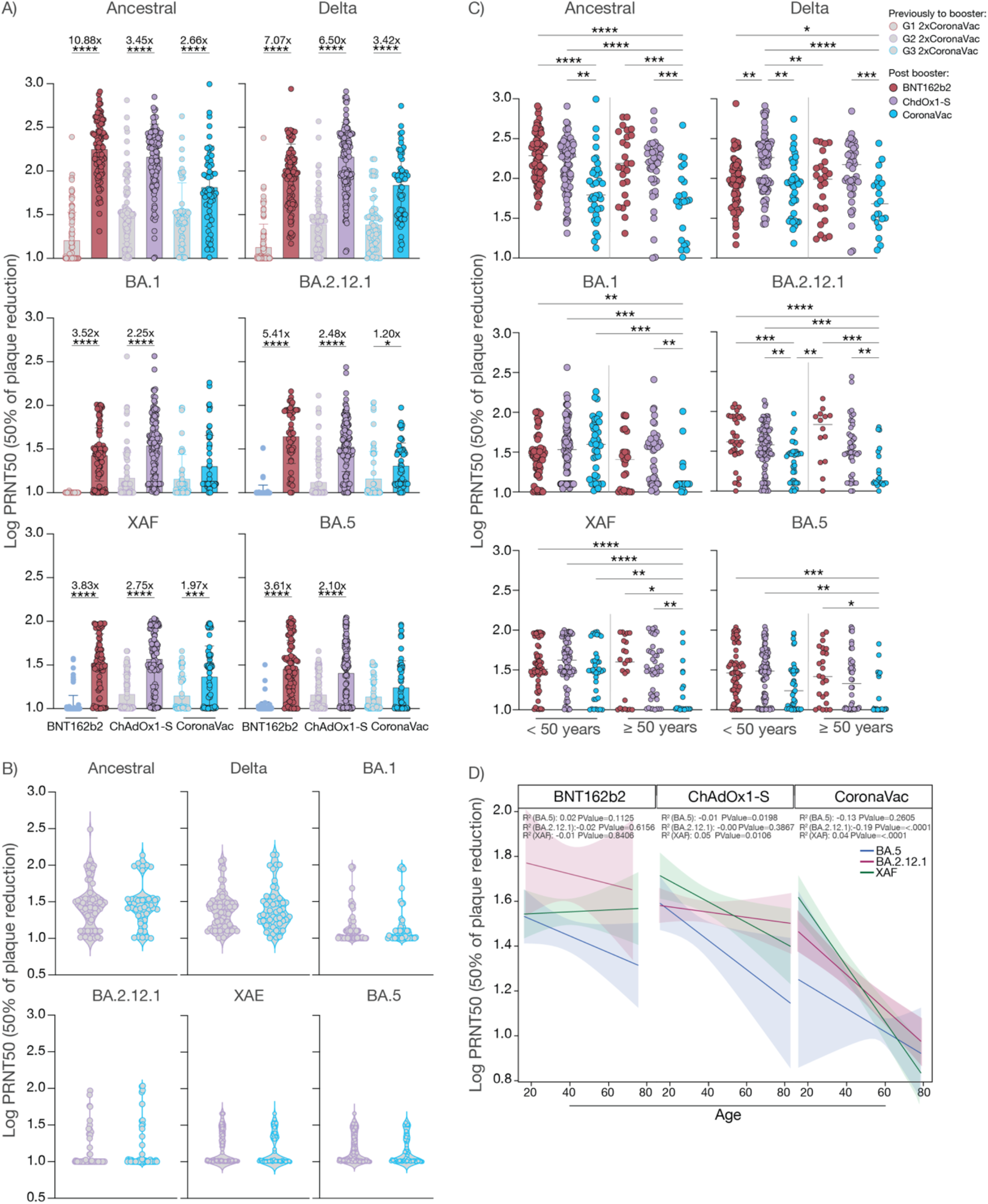
Characterization of neutralization response post heterologous versus homologous booster in fully vaccinated CoronaVac cohort. **a, b, c**, Plasma neutralization capacity against the ancestral strain (WA1, USA), Delta, BA.1, BA.2.12.1, XAF, and BA.5. Fully vaccinated CoronaVac participants received BNT162b2, ChAdOx1-S, or CoronaVac booster dose. Plasma samples were collected at baseline, prior to vaccination booster and 28 days after the booster dose. Participants were stratified based on the booster dose vaccine into three groups (G1, BNT162b2; G2, ChAdOx1-S; G3, CoronaVac). **a**, Plasma neutralization capacity comparisons at baseline and 28 days post booster dose. The median fold change values in antibody titer post booster vaccination are indicated. G1, n = 101; G2, n = 93; G3, n = 61. Total neutralization titers against: ancestral strain, Delta and BA.1 (BNT162b2, n = 101; ChAdOx1-S, n = 131; CoronaVac, n = 61); BA.2.12.1(BNT162b2, n = 49; ChAdOx1-S, n = 131; CoronaVac, n = 61); XAF, and BA.5 (BNT162b2, n = 86; ChAdOx1-S, n = 131; CoronaVac, n = 61). **b**, Plasma neutralization titers at baseline. **c**, Plasma neutralization capacity comparisons 28 days post booster dose by age groups. < 50 years, vaccinated participants aged 49 or younger; ≥ 50 years, vaccinated participants aged 50 years or older. Neutralization titers for < 50 years participants against: ancestral strain, Delta and BA.1 (BNT162b2, n = 76; ChAdOx1-S, n = 87; CoronaVac, n = 39); BA.2.12.1(BNT162b2, n = 35; ChAdOx1-S, n =87; CoronaVac, n = 39); XAF, and BA.5 (BNT162b2, n = 35; ChAdOx1-S, n = 87; CoronaVac, n = 39).Neutralization titers for ≥ 50 years participants against: ancestral strain, Delta and BA.1 (BNT162b2, n = 25; ChAdOx1-S, n = 44; CoronaVac, n = 22); BA.2.12.1(BNT162b2, n = 14; ChAdOx1-S, n =44; CoronaVac, n = 22); XAF, and BA.5 (BNT162b2, n = 25; ChAdOx1-S, n = 87; CoronaVac, n = 39). **d**, Correlation and linear regression comparisons of neutralizing titers against the current circulating VOCs XAF, BA.2.12.1, and BA.5 by age post vaccination booster. Regression lines are shown as green (XAF), red (BA.2.12.1) or blue (BA.5). Pearson’s correlation coefficients and linear regression significance are indicated, and shading represents 95% confidence interval. Significance was assessed by One-way ANOVA corrected for multiple comparisons using Tukey’s method. Each dot represents a single individual. Horizontal bars represent average ± s.d.****p < .0001 ***p < .001 **p < .01 *p < .05.

**Figure S3.**
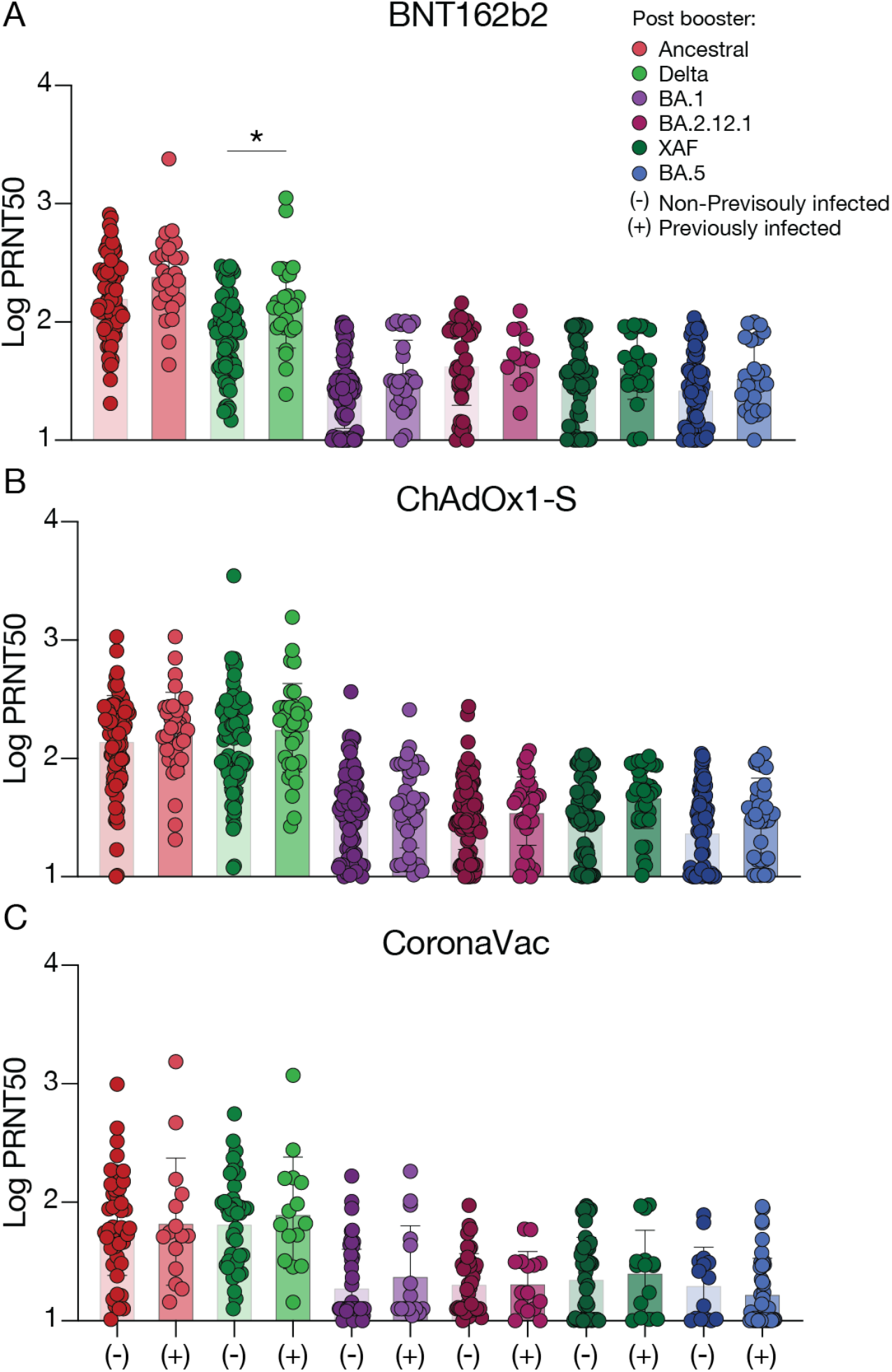
Comparison of neutralizing activity by SARS-CoV-2 infection status. **a**, Plasma neutralization capacity against the ancestral strain and variants of concern in fully CoronaVac vaccinated participants followed by booster dose with BNT162b2, ChAdOx1-S, or CoronaVac. Plasma neutralization titers measured at 28 days after the booster shot in non-previously infected (light color) and previously infected participants (dark color). Significance was measured using One-way ANOVA corrected using Tukey’s test. (–) Vaccinated–uninfected (BNT162b2, n = 75; ChAdOx1-S, n = 94; CoronaVac, n = 46); (+) vaccinated–previously infected (BNT162b2, n = 26; ChAdOx1-S, n = 37; CoronaVac, n = 15). Each dot represents a single individual. Horizontal bars represent average ± s.d. *p < .05.

**Table S1:** Detailed clinical data for each cohort. Clinical information, demographics and exact counts for immunological data.

| Individuals Variables | N | (%) | Mean | SD |
| --- | --- | --- | --- | --- |
| **Brazilian Cohort** |  |  |  |  |
| ChAdOx1-S booster |  |  |  |  |
| Age | 131 |  | 39.23 | 17.31 |
| < 50 years | 87 | 64.41 | 28.38 | 7.76 |
| ≥ 50 years | 44 | 33.59 | 60.68 | 8.75 |
| Gender |  |  |  |  |
| Male | 55 | 41.98 |  |  |
| Female | 76 | 58.01 |  |  |
| Infection Status |  |  |  |  |
| Non- Previously infected | 94 | 71.75 |  |  |
| Previously infected | 37 | 28.24 |  |  |
| Days since last infection |  |  | 321.16 | 173.57 |
| Days since last CoronaVac dose |  |  | 155.70 | 58.21 |
| CoronaVac booster |  |  |  |  |
| Age | 61 |  | 38.64 | 17.05 |
| < 50 years | 39 | 69.93 | 27.36 | 7.94 |
| ≥ 50 years | 22 | 36.06 | 58.64 | 7.89 |
| Gender |  |  |  |  |
| Male | 27 | 44.26 |  |  |
| Female | 34 | 55.74 |  |  |
| Infection Status |  |  |  |  |
| Non- Previously infected | 46 | 75.40 |  |  |
| Previously infected | 15 | 24.59 |  |  |
| Days since last infection |  |  | 369.13 | 196.48 |
| Days since last CoronaVac dose |  |  | 165.49 | 49.99 |
| **Dominican Republic Cohort** |  |  |  |  |
| BNT162b2 booster |  |  |  |  |
| Age | 101 |  | 40.4 | 13.4 |
| < 50 years | 76 | 75.24 | 33.96 | 8.27 |
| ≥ 50 years | 25 | 24.75 | 58.52 | 7.41 |
| Gender |  |  |  |  |
| Male | 31 | 30.7 |  |  |
| Female | 70 | 69.3 |  |  |
| Infection Status |  |  |  |  |
| Non- Previously infected | 75 | 74.25 |  |  |
| Previously infected | 26 | 25.74 |  |  |
| Days since last infection |  |  | 326.26 | 131.01 |
| Days since last CoronaVac dose | 101 |  | 113 | 33 |
| **Combined Cohorts** |  |  |  |  |
| Total Age | 293 |  | 39.38 | 15.95 |
| Total Gender |  |  |  |  |
| Male | 180 | 61.43 |  |  |
| Female | 113 | 38.56 |  |  |

## Notes

### Funding Statement

AstraZeneca Externally sponsored scientific research program: Brazilian clinical cohort set-up team.
Women Health Research at Yale Pilot Project Program: AI.
Fast Grant from Emergent Ventures at the Mercatus Center: AI and NDG.
Mathers Foundation, and the Ludwig Family Foundation, the Department of Internal Medicine at the Yale School of Medicine, Yale School of Public Health and the Beatrice Kleinberg Neuwirth Fund
Howard Hughes Medical Institute: AI.
CAPES-YALE fellowship: VSM.

### Author Declarations

Ethics statement: This study was approved by the National Research Bioethics Committee of Brazil (CONEP, CAAE 50457721.9.0000.0175) and National Bioethics Committee of the Dominican Republic (CONABIOS). The participants received two doses of the inactivated whole-virion vaccine CoronaVac followed by a single BNT162b2, ChAdOx-S, or CoronaVac booster dose. The interval between the second dose of CoronaVac and booster shot was at least four weeks. The Brazilian cohort received ChAdOx1-S and CoronaVac boosters, that were administrated between November 27, 2021 and February 03, 2022. The Dominican Republic received two doses of CoronaVac followed by the mRNA vaccine BNT162b2 booster, that was administrated between July 30 and August 27, 2021. Informed consent was obtained from all enrolled vaccinated individuals. None of the participants experienced serious adverse effects after vaccination.

